# Mapping Evidence on Road Safety Information Management Systems in Sub-Saharan Africa: Scoping Review Protocol

**DOI:** 10.1101/2024.02.13.24302651

**Authors:** Shemsedin Amme, Sheka Shemsi Seid, Mekonen Bogale, Demuma Amdisa, Abdusemed Husen, Getachew Tilahun, Nega Jibat, Getachew Mamo

**Author notes:** **Corresponding:** Shemsedin Amme Ibro.

## Abstract

**Background:** Road safety information management systems (RSIMS) play a vital role in improving road safety in sub-Saharan Africa (SSA) by providing reliable and timely data and information for evidence-based decision making. However, little is known about RSIMS in SSA.

**Objective:** The objective of this scoping review is to map the extent and type of evidence on RSIMS in Sub-Saharan Africa.

**Inclusion criteria:** This review will include sources that report on the application of RSIMS in SSA. The source of information that have considered the use of electronic data systems or software related to any aspects of the road safety management will be included irrespective of their designs or methods. Sources that focused on other populations, concepts, or context will be excluded.

**Methods:** A comprehensive search of published studies in electronic databases such as Scopus, PubMed, Embase, PubMed, RSKC, ARSO, Lens, and in the sources of grey literature will be carried out to identify relevant sources of information reported in English from 2019 onwards. A scoping review will be performed based on the JBI frameworks using Parsifal platform. Reviewers will independently screen the sources for eligibility, extract data using a JBI-adapted tool, analyze data using descriptive statistics and thematic analysis. The results will be presented in tables, figures, diagrams, and a narrative summary.

**Registration details:** This protocol for this scoping review has been registered on OSF, https://osf.io/6e2mx/.

## Introduction

Road traffic accidents (RTAs) are one of the major threats to humanity, casting a long shadow on the health and socioeconomic well-being of the global community[1]. According to the World Health Organization (WHO), in 2023, an estimated 1.2 million people died from RTAs, while 30–50 million suffered non-fatal injuries and permanent disabilities[2]. Low- and middle-income countries (LMICs) are disproportionately bearing the devastating human and economic impacts of RTAs, accounting for 93% of global deaths and costing 3% of their annual gross domestic product (GDP) [1,3].

Sub-Saharan Africa (SSA) represents the highest RTA fatality rates in the world, accounting for 19 deaths per 100,000 population[4], despite the lowest number of the world’s vehicles[5]. However, the burden of RTAs in SSA is underreported due to paucity of data[6]. Nevertheless, the burden is expected to increase by double more than a fold by 2030, due to rapid urbanization, population growth, motorization, and inadequate road infrastructure unless there is strong preventive measure[5]. In addition to the devastating human costs, SSA also bear the brunt of the RTA’s impacts on socioeconomic development, as the young and most productive parts of the society are disproportionately affected by road accidents[4]. Moreover, lack of strong and quality healthcare adds an additional layer of complexity to the problems, making RTA major public health problem in the region[6].

To address this soaring burden of RTAs, the WHO proposed safe system approach - a holistic strategy known as “Decade of Action for Road Safety 2021-2030”, across the world including in SSA[7]. Establishing data system is one of the strategies identified by this global decade of action, indicating the indispensable role of data [7]. The road safety data have crucial role in awareness raising, characterizing the accident, diagnosing its causes, and thereby to act as evidence and inform policy and program interventions, resource allocation, and monitoring and evaluating progress [8,9]. However, road safety data often relies on patchwork of data sources that include incomplete or inaccessible in SSA due to multifaceted challenges that hindering the understanding of the true magnitude of the problem, its consequences, decision making, and progress toward improvements [10–18].

Like other SSA countries, Ethiopia also faces a serious road safety problem, and one of the four countries accounting for more than half of the RTA fatalities in the region of SSA. According to the WHO report, in 2016, the estimated RTA fatalities was 27,326, with death rate 26.7 per 100000 population, and total of 409,890 serious non-fatal injuries. The report also indicates that the RTA cause devastating the economic loss in the country, as the 60% of the accidents occurring in the economically productive age groups (15 - 64 years), as well as causing more than $6.5 billion estimated total costs of fatalities and serious injuries[19]. The total annual GDP loses was estimated to be 8.8% of the GDP [19], which is more than twice of the country’s annual healthcare expenditure (4.2% of GDP) [19]. However, this WHO’s statistics variates with the official government’s, and Global burden of diseases (GBD) reports, which reported the road fatalities 4,352 and 9,639 for the same year, 2016, indicating limitations in data collection and dissemination[19]. Moreover, the safety data in the country’s rising the issues of quality and accuracy, as it relies on the manually processed paper works [20]. As a result, evidence have been indicating that the accurate burden of the RTA in Ethiopia is underreported and estimated to be at least double of what actually reported[13,20,21]. For instance, in their study Abegaz(2014) identified a significant underreporting and discrepancies between the police and hospital accident data in the country[20]. Ethiopia has taken some steps to address its road safety challenges including developing a national road safety strategy (2021-2030) [22], and slight reduction has been observed in the magnitude and death rates, while the aim is to reduce by 50% by 2030 [21–24]. Though, the country recognized the need for strengthening road safety data as one of priority areas, efforts yet remain to bring changes towards reliable and timely data and information management system [13,20].

Road safety data/information management system (RSIMS) is one of the key components of an effective road safety system suggested to overcome the challenges of poor road accident records and information management in each country across the world [25]. RSIMS is an electronic system that collects, stores, analyses and disseminates data and information on road safety issues [26–28]. Moreover, it plays essential roles in improving road safety through providing timely and evidence-based insight and information for decision-makers, practitioners, researchers, and the public[9,29]. Such systems can provide timely and accurate data that can serve several purposes including for awareness creation and advocacy, policy formulation, planning of program or project interventions, resource allocation, and monitoring and evaluation[9,27,29]. Despite the dire burden of RTA, gaps with data, and importance of RSIMS in improving road safety, there is a lack of comprehensive and systematic evidence on the current state, experience, and challenges with RSIMS implementation in SSA. As a result of this, road safety data issues and opportunities for improvement in most of the countries in the region of SSA, including in Ethiopia are not well understood or documented. Most of the previous studies are primary studies focused on the specific aspects of crash data such as sources, accuracy, quality, and geospatial distribution, while no review addressed entire spectrum of, and regional perspectives on the RSIMS issues and solutions exclusively in SSA[14,17,18,28,30–34].

Given this background, there is a need for a comprehensive and systematic overview of the literature on RSIMS in SSA, with a focus on Ethiopia. This will identify the evidence and knowledge gaps on RSIMS in the region, and inform future research, policy, and practice on road safety and RTA prevention. Therefore, this scoping review aims to fill this knowledge gap by mapping and synthesizing the existing literature on RSIMS in SSA, with an emphasis on Ethiopia. We will review both peer-reviewed and grey literature that address the technical, operational, or policy aspects of RSIMS in SSA, to examine the conceptualization, development, and implementation of RSIMS to improve road safety data and information in the region.

## Objective

The aim of this scoping review is to provide a comprehensive overview of the extent and state of knowledge on RSIMS in SSA. By systematically exploring and mapping the current state of knowledge on RSIMS, this review will inform further research and strategies for improving road safety outcomes in the region, with emphasis on Ethiopia.

## Research Questions

This scoping review aimed to answer this primary research question: What is the current state of evidence on RSIMS in SSA, and its practical and policy implications to Ethiopia?

Specifically, this scoping review will address the following four sub-research questions:

1. What are the core concepts in, characteristics, and components of RSIMS in SSA, and how are they implemented and utilized across different levels (e.g., accident scene, health facility, traffic police, national)?
2. What are the benefits, facilitators, and challenges for implementing RSIMS in SSA technological, institutional, data quality, and what factors influence their effectiveness and sustainability?
3. How RSIMS tailored to address the unique context of SSA countries (e.g., infrastructure, data collection, user needs), ensuring their relevance and effectiveness?
4. What are the best practices, lessons learned, and recommendations for improving RSIMS in SSA, and what are the implications for practice and policy in Ethiopia?

## Inclusion Criteria

The inclusion criteria for this scoping review are derived from research questions and objectives using a population, concept, and context (PCC) framework.

### Population

This review will include sources that report on or involve any road safety stakeholders including road users, healthcare providers, law enforcement officials, practitioners, researchers, and policymakers, to understand RSIMS from various perspectives and levels.

### Concept

Source will be included if it reported on the development or applications of electronic data systems or software related to any aspects of the road safety management. This includes any specific digital technology and applications relevant to RSIMS, aimed at data acquisition, storage, processing, retrieval, and dissemination of information on road safety. The source should report desired outcomes such as system components (including functionalities, data sources, incident information, indicators, platforms), addressing development and utilization process, as well as benefits, feasibility, and challenges of implementation. The review will not consider sources that not reporting on or addressing any of these aspects.

### Context

The sources that reporting on the utilization of RSIMS at any level, including accident scenes, health facilities, traffic police, and national/regional levels, and engaging stakeholders at each level will be included to ensure the comprehensive coverage of RSIMS implementation in different contexts. To be considered in the review, the source should report on or conducted in any SSA country or region, based on the on the World Bank definitions for low-income and lower-middle-income economies in SSA [35]. Sources that not specific to any country or region in SSA will be excluded, due to lack of generalizability of the findings to the contexts of the region.

### Source Type

To ensure broaden scope that capture diverse information sources, this review will consider both published and grey sources that reporting empirical or non-empirical evidence relevant to RSIMS, regardless of design or methodology they used. Specifically, primary studies including quantitative, qualitative, and mixed method studies are eligible sources of information. In addition, review articles including systematic and scoping reviews, as well as narrative, descriptive and explanatory literature reviews will also be included depending on the research question. Furthermore, reports, guidelines, conference proceedings, dissertations, informed perspectives, policy recommendations, texts and opinions papers will be considered for inclusion, if proving evidence relevant to the research questions. This review will exclude study protocols, opinion pieces, editorials, letters, news articles, and abstracts of articles that are not available in full texts as they do not provide sufficient information for this study.

The timeframe of including sources is that, if they published or reported in the last five years, after 2019 to ensure capturing relevant and current information. The source will be eligible if reported in English considering that English is a widely used research language in SSA and spoken by the research team.

## Methods and Analysis

This scoping will be guided by Johanna Briggs Institute (JBI) frameworks[36], Preferred Reporting Items for Systematic Reviews and Meta-analyses extension for scoping review (PRISMA-ScR) guideline[37] which are appropriate for conducting a systematic and transparent scoping review. The protocol for this review is registered on OSF[38]. This review will be commenced by March 2024 and accomplished within eight months. By implementing thorough and transparent methodology this review will addresses its proposed objectives.

### Search Strategy

A three-stage search strategy will be employed to locate relevant sources of information in electronic databases and sources of grey literature.

#### Stage 1

Database Search: Database search for published studies involve three steps involving a preliminary search of information, stage of the development of full search terms, and full search in targeted electronic databases. First, preliminary searches in limited databases including Scopus and PubMed will be done using free text search forms to identify relevant articles and abstracts. This will enable to identify relevant keywords and indexing terms, that inform the development initial comprehensive search strategy incorporating indexing terms such as “traffic accidents,” “road safety information systems,” and “Sub-Saharan Africa”. Moreover, developed initial comprehensive search strategy will be entered to selected databases, and PubReMiner platform [39], to understand the quality of search results, identify and characterize the commonly used keywords, their synonyms, and index terms relevant to the research questions.

The identified relevant keywords and indexing terms will be refined and combined using appropriate Boolean operators (AND/OR), and appropriate truncation (?/*). Then, developed search terms will be further evaluated for any syntactical errors, quality of search results, and diagrammatically to visually understand relationship between the concepts in the query and to further refine using online platforms including the Searchrefinery - SR Accelerator Tool [40], and 2DSearch platform [41]. Moreover, the sensitivity and specificity of the search strategy will be evaluated to ensure it capturing sources relevant to the research questions and excluding irrelevant studies as possible. The search strategy /queries for selected databases for supplemented to this protocol in the Appendix I.

Before the final search in the databases, full search strategy will be translated and piloted to each targeted database according to its syntax and functionality. Translation of the full search strategy to databases will be facilitated by online tools such as Polyglot SR Accelerator Tool[42], and 2DSearch platform[41], otherwise manually customized for databases not supported by these tools. The targeted databases to retrieve published studies will include Scopus, Embase, PubMed, The Lens, Transport Research International Documentation (TRID), Africa-Wide Information (AWI), WHO library, Road Safety Knowledge Centre (RSKC), African Journals Online (AJOL), African Road Safety Observatory (ARSO), European Road Safety Observatory (ERSO), and Using Digital Innovation and Technology to Advance Road Safety (UNITAR) (Table 1). Additional relevant databases will be considered based on the quality of the results, and research questions if required. Final full search of information in the targeted databases using a piloted and customized search strategy will be conducted. The language and publication year filters will be employed, for focusing on relevant, recent sources. In addition, advanced search functionalities in databases such as proximity operators, truncation wildcards, and field searching will be employed to refine results.

**Table 1:** Target databases, search providers, and links to search providers.

| Database | Search Provider | Link to Search Provider |
| --- | --- | --- |
| Scopus | Elsevier | https://www.scopus.com/ |
| EMBASE | Elsevier | https://www.embase.com/ |
| PubMed (MEDLINE) | National Library of Medicine (NLM) | https://pubmed.ncbi.nlm.nih.gov/ |
| The Lens | Cambia | https://www.lens.org/search/ |
| Semantic Scholar | Semanticscholar | https://www.semanticscholar.org/ |
| Transport Research International Documentation (TRID) | Transportation Research Board (TRB) | https://trid.trb.org/ |
| African Journals Online (AJOL) | African Journals OnLine (AJOL) | https://www.ajol.info/ |
| Africa-Wide Information (AWI) | EBSCOhost | https://www.ebsco.com/products/research-databases/africa-wide-information |
| Road Safety Knowledge Centre (RSKC) | Road Safety GB | https://roadsafetyknowledgecentre.org.uk/ |
| African Road Safety Observatory (ARSO) | African Union Commission (AUC) and partners | https://www.roadsafetyobservatory.africa/ |
| European Road Safety Observatory (ERSO) | European Commission | https://ec.europa.eu/transport/road_safety/specialist/erso_en |
| Using Digital Innovation and Technology to Advance Road Safety (UNITAR) | UNITAR | https://www.unitar.org/about/news-stories/news/using-digital-innovation-and-technology-advance-road-safety |
| WHO global health library | World Health Organization (WHO) | https://www.who.int/library/ |
| Research4Life/HINARI | Research4Life | https://portal.research4life.org/ |

#### Stage 2

Reference List Search: The reference lists of the included studies will be systematically searched using online platform Citation Chaser tool to identify relevant backward references and forward citations. This will be further supplemented by hand searches for potentially missed sources. In addition, databases like Connected Papers, Researchrabbit, ResearchGate, and Google Scholar Alerts for other relevant sources of evidence will be used for additional studies (Table 2).

**Table 2:** Sources of published and unpublished literature and link databases.

| Database/source | Link to databases |
| --- | --- |
| Google Scholar | https://scholar.google.com/ |
| EBSCO open dissertation | https://www.ebsco.com/products/research-databases/open-dissertations |
| Worldwidescience | https://www.worldwidescience.org/ |
| Researchrabbit | https://researchrabbit.ai/ |
| Connected Papers | https://www.connectedpapers.com/ |
| ResearchGate | https://www.researchgate.net/ |
| Google | https://www.google.com/ |

Moreover, the authors of the sources will be contacted to request the missing or additional data for clarification (unclear information), where required. The outcomes of the authors contact will be documented and reported in the review report.

#### Stage 3

Grey Literature Search: Electronic databases such as Google Scholar, EBSCO dissertation, and Worldwidescience will be used for unpublished studies, conference proceedings, thesis and dissertations. In addition, grey literature such as reports, policy recommendation, and government documents, will be identified through targeted searches of relevant websites and organizational repositories including World Road Safety Association (PIARC), World Health Organization, World Bank, African Development Bank, United Nations Economic Commission for Africa, and Sub-Saharan Africa Transport Policy Program. Language and Publication Year restriction will be considered based on the inclusion criteria.

### Selection of Sources

Following the search, full citations of identified sources will be imported to and collated using Zotero version 6.0.30 (Corporation for Digital Scholarship, Virginia, USA)[43]. This will be followed by deduplication using online platform the Deduplicator SR-Accelerator Too[44]. A two-stage screening process will be employed to select eligible sources against inclusion criteria, to ensure objectivity and minimizes bias. First, title and abstract screening will be done by importing the deduplicated citations of identified sources to The Screnator SR-Accelerator Tool [44], a web platform for screening. This will be done by two independent reviewers through assessing and screening against the inclusion criteria.

Next, full-text of the potentially eligible sources will be retrieved, and their citation details imported into the Parsifal review platform - a web-based tool for systematic reviews in software engineering[45]. Then, full text will be assessed in detail against the inclusion criteria by two or more independent reviewers. For the sources excluded at a full text screening stage, reasons for exclusion will be recorded and reported in the scoping review for transparency. Any disagreements between reviewers at each stage of screening will be resolved through discussion or involving a third reviewer.

The results of the search and screening process will be documented and reported in full scoping review, and presented using the Preferred Reporting Items for Systematic Reviews and Meta-analyses extension for scoping review (PRISMA-ScR) flow diagram[37].

### Data Extraction

Data extraction will be done using a JBI guideline-adapted tool [36], on Parsifal review platform[45], by independent reviewers. Extracted data will include specific details about the bibliographic information, population, concept, context, methods, and key findings related to the research questions from the included articles. We adapted the JBI data extraction tool [36] for scoping reviews to suit the specific objectives and questions of our review by adding the one data field for implications /recommendations, while modifying one field on the intervention characteristics and comparison groups, as concept/intervention. Detailed data extraction tool fields and definitions supplemented in Appendix II of this protocol.

The data extraction fields will include the following categorized structured coding scheme:

Specific data fields and categories will be further defined based on the research questions and anticipated themes to ensure targeted data collection achieved. The data extraction tool will be piloted by the reviewers on sample of sources before actual data extraction. Modification and revision will be done as required based on pilot test.

Data extraction fields will be coded to online review Parsifal platform[45], to facilitate efficient and standardized data collection. The data and information extracted from each source will be assessed for accuracy and validated against the original source, and any errors or discrepancies corrected or clarified as needed. Any disagreements that arise between the reviewers will be resolved through discussion, or with an additional third reviewer. In addition, authors of papers will be contacted to request any missing or additional data, where required.

The quality assessment will not be formally assessed, as this is not a requirement for scoping reviews. However, any limitations or strengths of the sources that may affect the interpretation or generalization of the findings will be acknowledged and discussed in the review.

### Data Synthesis and Presentation

Extracted data will be analysed using descriptive statistics and thematic analysis to summarize and synthesize information extracted from the included sources. The descriptive statistics will be used to provide summarized information overview of the characteristics and distribution of the sources and their findings according to key variables such as year of publication, country, type, and design. The thematic analysis will be used to identify the code schemes developed in the data extraction stage, and organize them using the key themes and subthemes that emerge from the data, and according to the research questions on Microsoft office (Microsoft Office, Microsoft, Redmond, Washington USA), using DocTools Add-ins (DoocTools ExtractData 1.5) [46]. Inter-rater variability of the codes and themes will be assessed ensure reliability.

Findings will be presented in a clear and concise manner, utilizing tables, figures, diagrams, and a comprehensive narrative summary that clearly addresses the research questions and provides actionable insights. This will include a table summarizing the characteristics and findings of each source, a figure showing the PRISMA flow diagram of the screening and selection process, and a diagram depicting the thematic map of the key themes and categories from the thematic analysis. Moreover, a narrative summary will be used to present and discuss the findings, providing a comprehensive and coherent description and interpretation of the results. This will be organized and presented using the themes including type and key components; the context and implementation; benefits and effects; challenges and facilitators; and the recommendations and implications to provide holistic understanding of the current state of knowledge on RSIMS in SSA.

## Dissemination Plan

The report of this review will be prepared following the structure and content recommended by the JBI Manual for scoping review[36] and the PRISMA-ScR reporting guidelines [37]. The finding will be disseminated to relevant stakeholders, including policymakers, researchers, practitioners, and communities using different channels such as publication on a peer-reviewed journals and presentation on the scientific conference that focuses on road safety or public health in Africa. Moreover, the findings will be used to guide the future studies, where this scoping of the review is part of the evidence and knowledge mapping.

## Expected Outcomes

This scoping review will contribute significantly to the ongoing efforts to improve RSIMS by providing a comprehensive overview of the current state of evidence on RSIMS in SSA, identifying the key gaps and challenges in its implementation and utilization, map potential solutions and recommendations for improvements, and inform practical and policy strategies for enhancing road safety in Ethiopia and other SSA countries.

## Data Availability

No datasets were generated or analysed during the current study. All relevant data from this study will be made available upon study completion.

## Funding

No funds were received for this review.

## Author’s contributions

All authors contributed equally to conceiving and writing this protocol. Authors SAI, SSS, MB, and GM developed the research questions and inclusion criteria of this protocol. SAI, NJ, and SSS designed the search strategy, and study selection plan, while AH, GT, and DA designed and tested data extraction tool, and data analysis and presentation plan.

## Competing of interest

The authors have declared that no competing interests exist.

## Notes

### Competing Interest Statement

The authors have declared no competing interest.

### Funding Statement

The author(s) received no specific funding for this work.

