## Supplemental files for "Mapping Evidence on Road Safety Information Management Systems in Sub-Saharan Africa: Scoping Review Protocol"

### Appendices

#### Appendix I: Search strategy

**Search terms: January 25, 2024**

##### PubMed:

1. Search: ((("accidents, traffic"[Mesh]) OR ("road safety"[tiab] OR "transport safety"[tiab])) AND ("Electronic Data Processing"[Mesh] OR "Information Management"[Mesh] OR "Health Information Management"[Mesh] OR "Integrated Advanced Information Management Systems"[Mesh] OR "Information Sources"[Mesh] OR "Information Dissemination"[Mesh] OR "Health Information Exchange"[Mesh] OR "Health Information Systems"[Mesh] OR "Geographic Information Systems"[Mesh] OR "Access to Information"[Mesh] OR "Information Storage and Retrieval"[Mesh] OR "Information Systems"[Mesh] OR "Health Information Interoperability"[Mesh] OR "Information Technology"[Mesh] OR "Geographic Mapping"[Mesh] OR "Electronic Health Records"[Mesh] OR Documentation[Mesh] OR "Online Systems"[Mesh] OR "Public Health Surveillance"[Mesh] OR "Spatial Analysis"[Mesh] OR "Data Mining"[Mesh])) AND (("africa south of the sahara"[Mesh]))
2. Filter: 2019 – 2024 [Publication Year] AND English[Language]

##### Scopus:

1. Search: (((INDEXTERMS("accidents, traffic")) OR (TITLE-ABS("road safety") OR TITLE-ABS("transport safety"))) AND (INDEXTERMS("Electronic Data Processing") OR INDEXTERMS("Information Management") OR INDEXTERMS("Health Information Management") OR INDEXTERMS("Integrated Advanced Information Management Systems") OR INDEXTERMS("Information Sources") OR INDEXTERMS("Information Dissemination") OR INDEXTERMS("Health Information Exchange") OR INDEXTERMS("Health Information Systems") OR INDEXTERMS("Geographic Information Systems") OR INDEXTERMS("Access to Information") OR INDEXTERMS("Information Storage and Retrieval") OR INDEXTERMS("Information Systems") OR INDEXTERMS("Health Information Interoperability") OR INDEXTERMS("Information Technology") OR INDEXTERMS("Geographic Mapping") OR INDEXTERMS("Electronic Health Records") OR INDEXTERMS(Documentation) OR INDEXTERMS("Online Systems") OR INDEXTERMS("Public Health Surveillance") OR INDEXTERMS("Spatial Analysis") OR INDEXTERMS("Data Mining"))) AND ((INDEXTERMS("africa south of the sahara")))
2. Filter: PY AFTER 2019 AND English [Language]

##### Embase:

1. Search: ((('accidents, traffic'/exp) OR ('road safety':ti,ab OR 'transport safety':ti,ab)) AND ('Electronic Data Processing'/exp OR 'Information Management'/exp OR 'Health Information Management'/exp OR 'Integrated Advanced Information Management Systems'/exp OR 'Information Sources'/exp OR 'Information Dissemination'/exp OR 'Health Information Exchange'/exp OR 'Health Information Systems'/exp OR 'Geographic Information Systems'/exp OR 'Access to Information'/exp OR 'Information Storage and Retrieval'/exp OR 'Information Systems'/exp OR 'Health Information Interoperability'/exp OR 'Information Technology'/exp OR 'Geographic Mapping'/exp OR 'Electronic Health Records'/exp OR Documentation/exp OR 'Online Systems'/exp OR 'Public Health Surveillance'/exp OR 'Spatial Analysis'/exp OR 'Data Mining'/exp)) AND (('africa south of the sahara'/exp))
2. Filter: 2019 – 2024 [Publication Year] AND English [Language]

#### Appendix II: Data extraction tool

Table 3: Data extraction fields and definitions

| **Field** | **Definition/Details** |
| --- | --- |
| Bibliographic details: | Author, Year, Title, Journal/Source name |
| Source type: | Journal article, review, report, guideline, thesis, etc. |
| Country: | The country or countries where the source conducted or focused on. |
| Population: | The stakeholders involved in or affected by the RSIMS including policy makers, healthcare providers, officials, practitioners, researchers, and road users. |
| Context/Setting: | Setting/context where the RSIMS was implemented (national, regional, local, health facility, police, scene), description of the resource availability, system characteristics, stakeholder involvement, and strategies used to adapt to the different contexts. |
| Methods: | The design or methodology used by the source to describe or evaluate the RSIMS such as quantitative, qualitative, mixed methods, or systematic/scoping review. |
| Concept/Intervention/: | The concepts in and characteristics of RSIMS implemented/evaluated including the definition or how the source defined or conceptualized RSIMS, their type, components, functions, and features, as well as level of the degree and type of implementation/interventions done or evaluated using a suitable framework or typology. |
| Outcome(s): | The main outcome(s) measured or reported by the sources. |
| Result(s): | The summary of the key finding(s) reported for the outcome measured or reported including the benefits, feasibility, challenges, and gaps of the RSIMS, as well as potential solutions or best experience/recommendations to overcome the challenges. |
| Conclusion(s): | The main conclusion(s) drawn by the authors based on the finding(s), such as conclusion, recommendations, and limitations, of the result(s). |
| Implications: | The main implications or recommendations for policy and practice in Ethiopia and broader SSA based on the findings and conclusions of the source in relation to the RSIMS. |

### Appendices

#### Appendix I: Search strategy

**Search terms: January 25, 2024**

##### PubMed:

3. Search: ((("accidents, traffic"[Mesh]) OR ("road safety"[tiab] OR "transport safety"[tiab])) AND ("Electronic Data Processing"[Mesh] OR "Information Management"[Mesh] OR "Health Information Management"[Mesh] OR "Integrated Advanced Information Management Systems"[Mesh] OR "Information Sources"[Mesh] OR "Information Dissemination"[Mesh] OR "Health Information Exchange"[Mesh] OR "Health Information Systems"[Mesh] OR "Geographic Information Systems"[Mesh] OR "Access to Information"[Mesh] OR "Information Storage and Retrieval"[Mesh] OR "Information Systems"[Mesh] OR "Health Information Interoperability"[Mesh] OR "Information Technology"[Mesh] OR "Geographic Mapping"[Mesh] OR "Electronic Health Records"[Mesh] OR Documentation[Mesh] OR "Online Systems"[Mesh] OR "Public Health Surveillance"[Mesh] OR "Spatial Analysis"[Mesh] OR "Data Mining"[Mesh])) AND (("africa south of the sahara"[Mesh]))
4. Filter: 2019 – 2024 [Publication Year] AND English[Language]

##### Scopus:

3. Search: (((INDEXTERMS("accidents, traffic")) OR (TITLE-ABS("road safety") OR TITLE-ABS("transport safety"))) AND (INDEXTERMS("Electronic Data Processing") OR INDEXTERMS("Information Management") OR INDEXTERMS("Health Information Management") OR INDEXTERMS("Integrated Advanced Information Management Systems") OR INDEXTERMS("Information Sources") OR INDEXTERMS("Information Dissemination") OR INDEXTERMS("Health Information Exchange") OR INDEXTERMS("Health Information Systems") OR INDEXTERMS("Geographic Information Systems") OR INDEXTERMS("Access to Information") OR INDEXTERMS("Information Storage and Retrieval") OR INDEXTERMS("Information Systems") OR INDEXTERMS("Health Information Interoperability") OR INDEXTERMS("Information Technology") OR INDEXTERMS("Geographic Mapping") OR INDEXTERMS("Electronic Health Records") OR INDEXTERMS(Documentation) OR INDEXTERMS("Online Systems") OR INDEXTERMS("Public Health Surveillance") OR INDEXTERMS("Spatial Analysis") OR INDEXTERMS("Data Mining"))) AND ((INDEXTERMS("africa south of the sahara")))
4. Filter: PY AFTER 2019 AND English [Language]

##### Embase:

3. Search: ((('accidents, traffic'/exp) OR ('road safety':ti,ab OR 'transport safety':ti,ab)) AND ('Electronic Data Processing'/exp OR 'Information Management'/exp OR 'Health Information Management'/exp OR 'Integrated Advanced Information Management Systems'/exp OR 'Information Sources'/exp OR 'Information Dissemination'/exp OR 'Health Information Exchange'/exp OR 'Health Information Systems'/exp OR 'Geographic Information Systems'/exp OR 'Access to Information'/exp OR 'Information Storage and Retrieval'/exp OR 'Information Systems'/exp OR 'Health Information Interoperability'/exp OR 'Information Technology'/exp OR 'Geographic Mapping'/exp OR 'Electronic Health Records'/exp OR Documentation/exp OR 'Online Systems'/exp OR 'Public Health Surveillance'/exp OR 'Spatial Analysis'/exp OR 'Data Mining'/exp)) AND (('africa south of the sahara'/exp))
4. Filter: 2019 – 2024 [Publication Year] AND English [Language]

#### Appendix II: Data extraction tool

Table 3: Data extraction fields and definitions

| **Field** | **Definition/Details** |
| --- | --- |
| Bibliographic details: | Author, Year, Title, Journal/Source name |
| Source type: | Journal article, review, report, guideline, thesis, etc. |
| Country: | The country or countries where the source conducted or focused on. |
| Population: | The stakeholders involved in or affected by the RSIMS including policy makers, healthcare providers, officials, practitioners, researchers, and road users. |
| Context/Setting: | Setting/context where the RSIMS was implemented (national, regional, local, health facility, police, scene), description of the resource availability, system characteristics, stakeholder involvement, and strategies used to adapt to the different contexts. |
| Methods: | The design or methodology used by the source to describe or evaluate the RSIMS such as quantitative, qualitative, mixed methods, or systematic/scoping review. |
| Concept/Intervention/: | The concepts in and characteristics of RSIMS implemented/evaluated including the definition or how the source defined or conceptualized RSIMS, their type, components, functions, and features, as well as level of the degree and type of implementation/interventions done or evaluated using a suitable framework or typology. |
| Outcome(s): | The main outcome(s) measured or reported by the sources. |
| Result(s): | The summary of the key finding(s) reported for the outcome measured or reported including the benefits, feasibility, challenges, and gaps of the RSIMS, as well as potential solutions or best experience/recommendations to overcome the challenges. |
| Conclusion(s): | The main conclusion(s) drawn by the authors based on the finding(s), such as conclusion, recommendations, and limitations, of the result(s). |
| Implications: | The main implications or recommendations for policy and practice in Ethiopia and broader SSA based on the findings and conclusions of the source in relation to the RSIMS. |
